# An internet-based survey of people with chronic lymphocytic leukemia in the UK

**DOI:** 10.1101/2025.08.05.25333089

**Authors:** Hilary Lindsay, Debbie Yates, Lelia Duley

## Abstract

**Background:** Chronic lymphocytic leukemia (CLL) is a common adult blood cancer. There is no cure, but a third of people never require treatment and new targeted drugs have improved outcome.

Understanding the experience of living with CLL is important for clinicians, health care providers and researchers.

**Methods:** This was an open internet-based survey of people with CLL, conducted by people with CLL. It was advertised via email and social media.

**Findings:** 1,009 people completed the survey. Respondents were more likely to be women (526, 52% female; 482, 48% male) and were younger at diagnosis (574, 57% <71 years) than the general CLL population.

For one third of respondents diagnosis was in the last five years. For 617 (61%) diagnosis was at a GP visit for another heath condition. 40% (406) reported no other health conditions. 558 (55%) respondents were currently on active monitoring, 261 (26%) were receiving treatment, and 190 (19%) in remission. Satisfaction with the speed of diagnosis, explanation of their CLL diagnosis and of the next steps in treatment was high.

A quarter of respondents reported no current signs and symptoms of CLL, or side effects of treatment. Fatigue was reported by 578 (57%), three quarters of whom experienced this daily. Fatigue was reported to have a high impact on day-to-day life. Other common problems were bleeding and bruising more than normal (263, 26%) and getting infections often (228, 23%).

From an emotional and a physical perspective, respondents reported their CLL had a higher impact on quality of life now than before the COVID-19 pandemic. Most respondents (829, 82%) received their treatment or monitoring at a hospital, for 253 (31%) contact was ‘mostly/all remote’.

**Conclusions:** Fatigue is a key problem for people with CLL. CLL impacts on the physical and emotional aspects of quality of life.

## INTRODUCTION

Chronic lymphocytic leukemia (CLL) is the most common adult blood cancer in the UK.[1] Each year around 4,720 people in the UK hear for the first time that they have CLL.[2] It is more common in men than women, roughly two thirds of those with CLL are men and one third are women. Diagnosis is often at older ages, with median age at diagnosis 72 years.[3,4] An estimated 33,570 people who are currently alive in the UK will have been diagnosed with CLL over the last 10 years.[2] At the time of diagnosis more than three quarters are asymptomatic and do not require immediate treatment. About one third never require treatment and remain on active monitoring (also known as ‘watch and wait’).[4] There is currently no cure for CLL, although modern targeted treatments have improved outcome substantially compared with chemoimmunotherapy.[5–7] Those with CLL may undergo a series of treatments with periods of remission between.

As treatment improves, a growing number of people are living with CLL. Hence, understanding the experiences of those living with this condition is increasingly important. For example, to help plan services and support, and to inform the prioritisation and design of research relevant to the needs of those with CLL. Also, people with CLL value a shared understanding of their experiences.

CLL and small lymphocytic lymphoma (SLL) are different forms of the same illness. When the blood cancer is mostly in the blood and bone marrow it is called CLL, and when it is mostly in the lymph nodes it is called SLL.[8]

This survey was conducted by the CLL Support Association UK, a small volunteer-led charity. The aim was to improve our understanding of the experiences and concerns of those living with by CLL in the UK.

## METHODS

This was an online survey of people with CLL or SLL. Partners of those with CLL or SLL could also respond, but their data are not included here. Reporting is consistent with the Checklist for Reporting Results of Internet E-Surveys CHERRIES guideline.[9] The CHERRIES checklist is included in supplementary file 1. We used convenience sampling to contact potential respondents. A link to the questionnaire was sent to everyone on the CLL Support newsletter email list. It was also posted in our Under 60’s WhatsApp Group and on our social media channels, including Facebook, Instagram and X. The questionnaire, data analysis and dissemination of results were largely based on the first CLL Support survey conducted in 2022.[10] The 2022 survey was therefore regarded as the protocol for this 2024 survey.

Research ethics committee review was not required, as the survey did not meet the criteria for such review in any of the four nations in the UK. Having clicked on the link to the questionnaire, it opened with a short introduction explaining the purpose of the survey. This introduction also explained that individual responses would be anonymous and confidential (see supplementary file 2). Respondents were then asked whether they agreed to take part on this basis. If they responded ‘yes’ this was considered consent and allowed access to the questionnaire. Completing the questionnaire was entirely voluntary, and respondents could exit the survey at any time.

### Questionnaire

A copy of the questionnaire is included in supplementary file 2. The online survey was tested by HL and DY. Slight amendments were made to wording to improve clarity. All the different pathways through the survey were checked to ensure people were asked the correct questions based on earlier answers. The information for respondents stated that they could drop out at any time. If people dropped out and subsequently rejoined, the survey took them back to the beginning.

### Data management and analysis

As for the 2022 survey, data management was by a market research company, Research Interactive Ltd (see https://research-interactive.com/). Research Interactive provided the dataset used for the analyses presented here. All analyses for this paper were conducted by the authors.

### Patient and Public Involvement

The survey was designed and conducted by members of CLL Support, a small volunteer-led charity. Therefore, people with CLL decided what questions were included in the survey, they tested the online questionnaire, they prepared and sent information about taking part in the survey to the CLL community, and they raised awareness of the opportunity to complete the survey via social media and the charity’s newsletter. Also, the 2022 questionnaire, on which our questionnaire was based, had been tested by people with CLL who were asked to check the questions were clear and that the questionnaire length was appropriate. For this paper, the analysis was planned and conducted by people with CLL who also wrote the paper.

## RESULTS

The survey opened on 4 March 2024 and closed on 15 April 2024. Several reminders to complete the survey were issued during this period. Overall, the survey link was opened 1,750 times. Of those who opened the link 1,274 completed the question about whether they agreed to take part and 1,247 completed information about a diagnosis of CLL or SLL. As the link potentially could be accessed multiple times by the same person, incomplete responses may have been duplicates. Therefore, data are reported only for the 1,110 respondents who reached the end of the survey. Most (956, 86%) responded via the link sent by email.

1,009 respondents said they had CLL or SLL (976 CLL, 33 SLL). The remaining 101 were a partner, carer or close friend of someone with CLL or SLL (98 partner or other family member, 3 carer or close friend). As the numbers are small, data for this group are not reported here.

People with CLL or SLL who completed our survey were more likely to be women (526, 52% female; 482, 48% male) and were younger at diagnosis (574, 57% <71 years) than the general population with CLL (table 1). Almost all respondents reported their ethnicity as white (983, 97%). Most (850, 84%) lived in England (table 1).

**Table 1.**
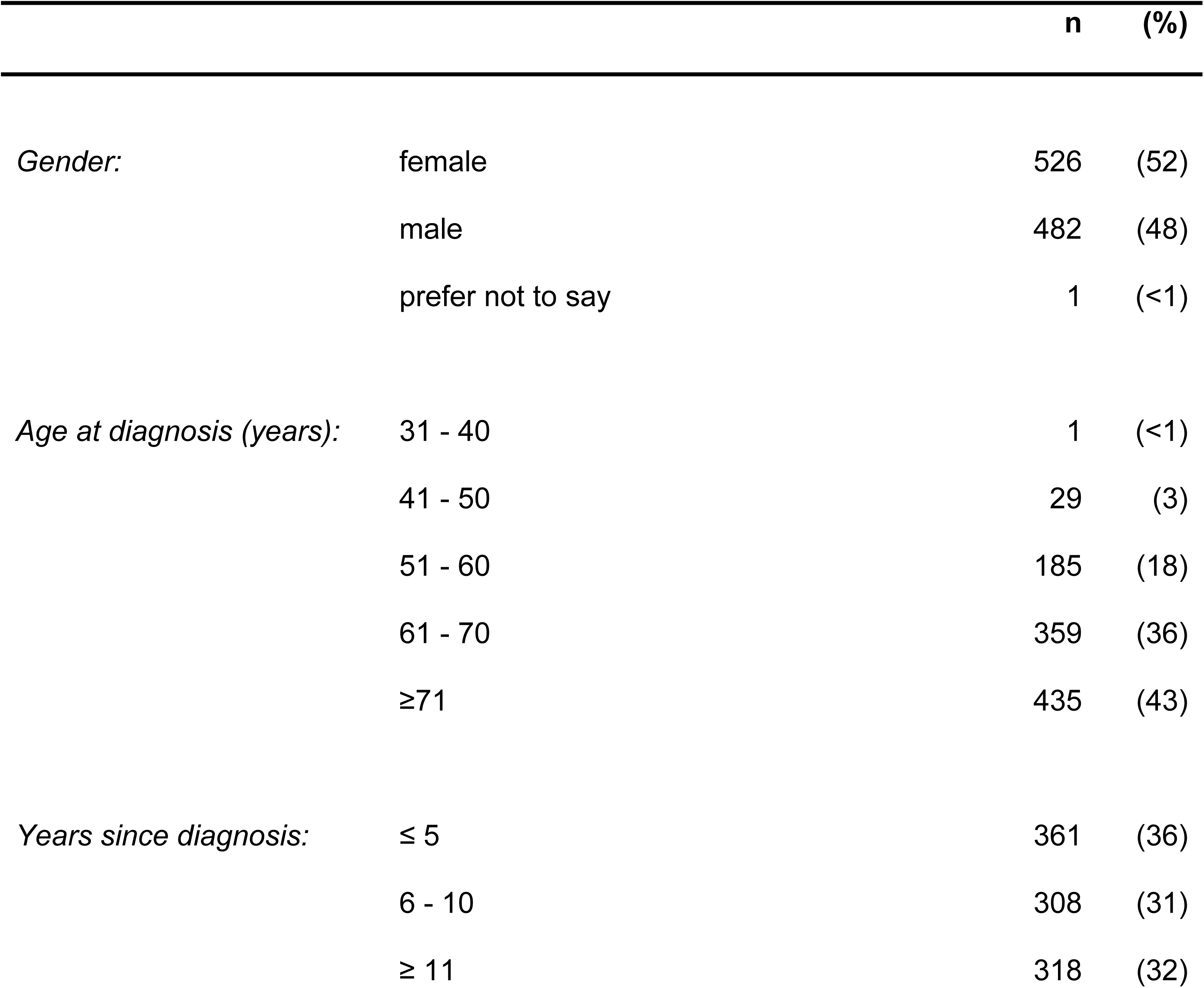
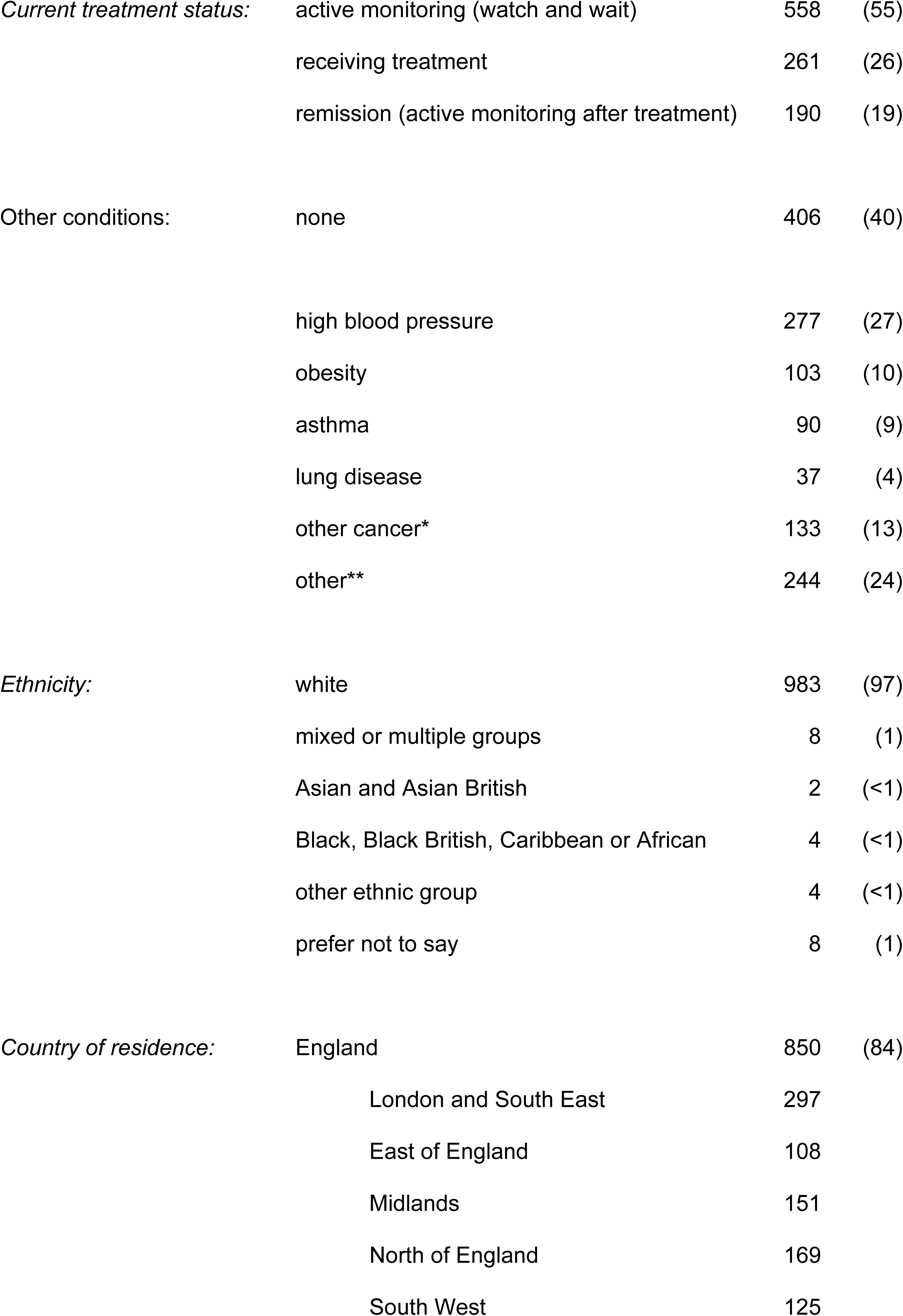
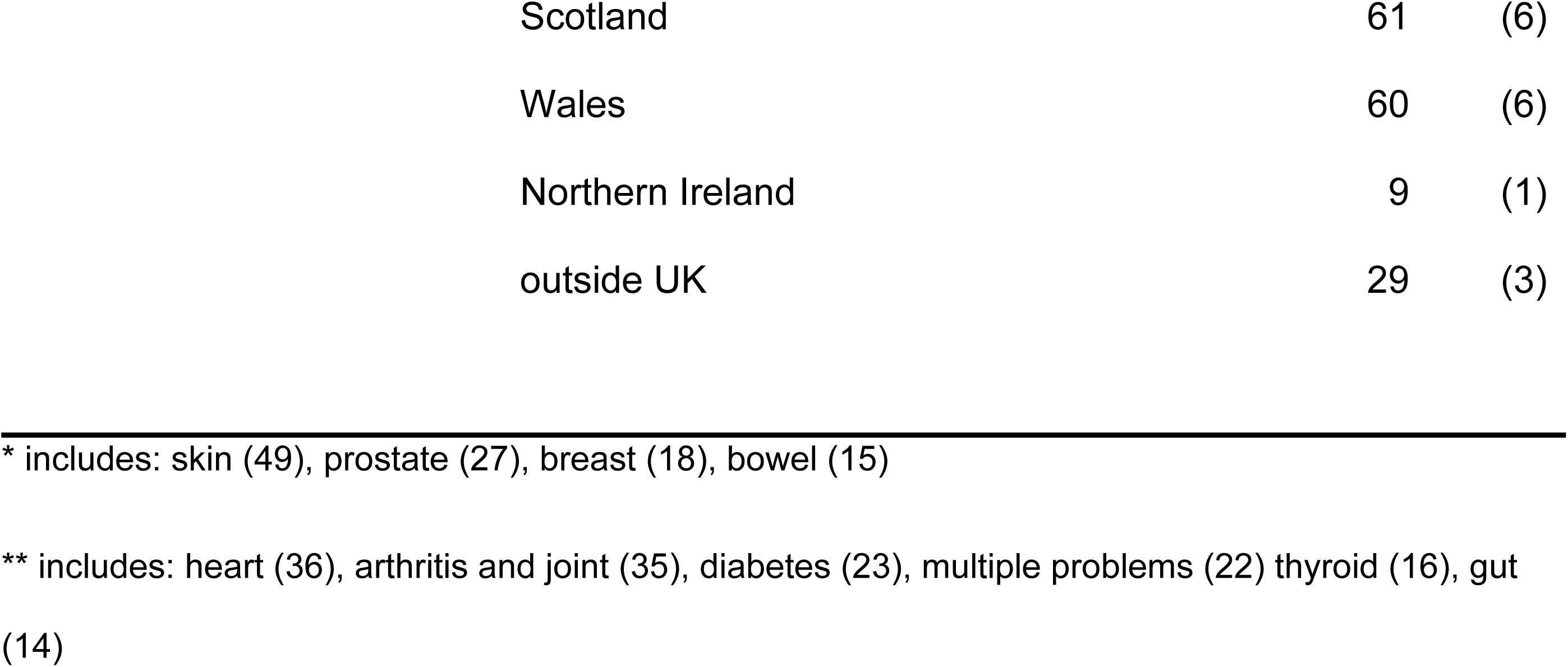
Demographic characteristics for the 1,009 respondents with CLL or SLL.

|  |  | n | (%) |
| --- | --- | --- | --- |
| *Gender:* | female | 526 | (52) |
|  | male | 482 | (48) |
|  | prefer not to say | 1 | (<1) |
| *Age at diagnosis (years):* | 31 - 40 | 1 | (<1) |
|  | 41 - 50 | 29 | (3) |
|  | 51 - 60 | 185 | (18) |
|  | 61 - 70 | 359 | (36) |
|  | ≥71 | 435 | (43) |
| *Years since diagnosis:* | ≤ 5 | 361 | (36) |
|  | 6 - 10 | 308 | (31) |
|  | ≥ 11 | 318 | (32) |
| *Current treatment status:* | active monitoring (watch and wait) | 558 | (55) |
|  | receiving treatment | 261 | (26) |
|  | remission (active monitoring after treatment) | 190 | (19) |
| Other conditions: | none | 406 | (40) |
|  | high blood pressure | 277 | (27) |
|  | obesity | 103 | (10) |
|  | asthma | 90 | (9) |
|  | lung disease | 37 | (4) |
|  | other cancer* | 133 | (13) |
|  | other** | 244 | (24) |
| *Ethnicity:* | white | 983 | (97) |
|  | mixed or multiple groups | 8 | (1) |
|  | Asian and Asian British | 2 | (<1) |
|  | Black, Black British, Caribbean or African | 4 | (<1) |
|  | other ethnic group | 4 | (<1) |
|  | prefer not to say | 8 | (1) |
| *Country of residence:* | England | 850 | (84) |
|  | London and South East | 297 |  |
|  | East of England | 108 |  |
|  | Midlands | 151 |  |
|  | North of England | 169 |  |
|  | South West | 125 |  |

|  |  |  |
| --- | --- | --- |
| Scotland | 61 | (6) |
| Wales | 60 | (6) |
| Northern Ireland | 9 | (1) |
| outside UK | 29 | (3) |

\* includes: skin (49), prostate (27), breast (18), bowel (15)
\*\* includes: heart (36), arthritis and joint (35), diabetes (23), multiple problems (22) thyroid (16), gut (14)

For just over one third of respondents their diagnosis was in the last five years (table 1). For almost two thirds (617, 61%) diagnosis was at a routine GP visit for some other heath condition (data not shown). Some (158, 16%) were seen by a GP with signs or symptoms of CLL. Largely, satisfaction with the speed of diagnosis, explanation of their CLL diagnosis and explanation of the next steps in treatment was high (figure 1). Over half of the respondents (558, 55%) were on active monitoring (also known as watch and wait) at the time they completed the survey, a quarter (261, 26%) were receiving treatment for their CLL, and one fifth (190, 19%) were in remission (table 1). Overall, 40% (406) reported no other health conditions.

**Figure 1.**
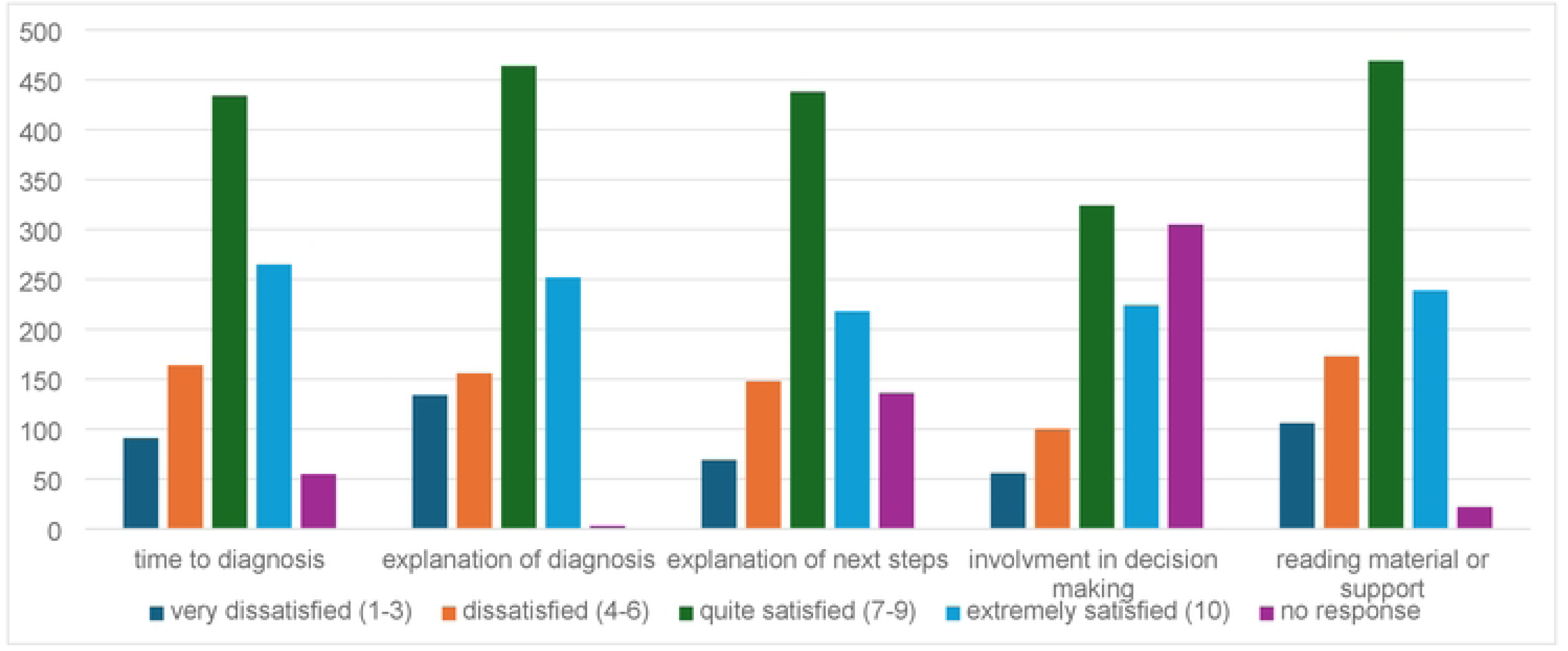
Responses to “how satisfied were you with the following aspects of CLL/SLL diagnosis and explanation?” (scale where 1= totally dissatisfied, 10= extremely satisfied)

A quarter of respondents reported no current signs and symptoms of CLL or side effects of treatment (table 2). Of those who did report these, many reported more than one. Fatigue was by far the most common symptom, reported by more than half (578, 57%) of respondents (table 2). Nearly three-quarters of those with fatigue said they experienced this every day (table 2). They also reported that fatigue had a high impact on their day-to-day life (figure 2). Just over a quarter of respondents reported bleeding and bruising more often than normal (263, 26%) and slightly less than a quarter that they were getting infections often (228, 23%) (table 2). The frequency of bleeding and bruising was relatively evenly distributed from daily to monthly and less often (table 2). However, the impact of this on day-to-day life was low to moderate (figure 2). Getting infections more often was largely reported as monthly or less often, but with a higher impact on day-to-day life. Although about one-fifth of respondents reported swollen glands, and for two-thirds this was daily, impact of this on day-to-day life was low to moderate. Other problems were reported less often (table 2).

**Figure 2.**
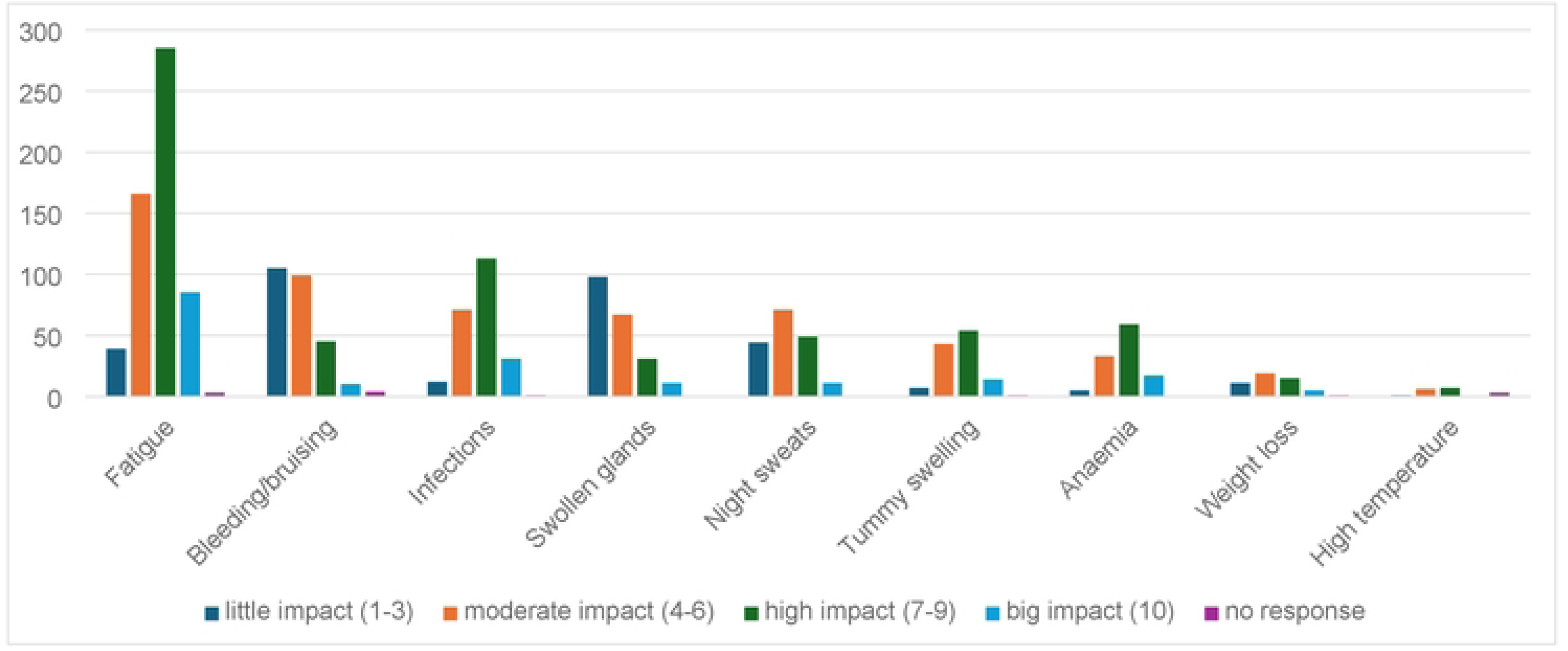
Impact of symptoms on day-to-day life (scale with 1= no impact, 10= a big impact)

**Table 2.**
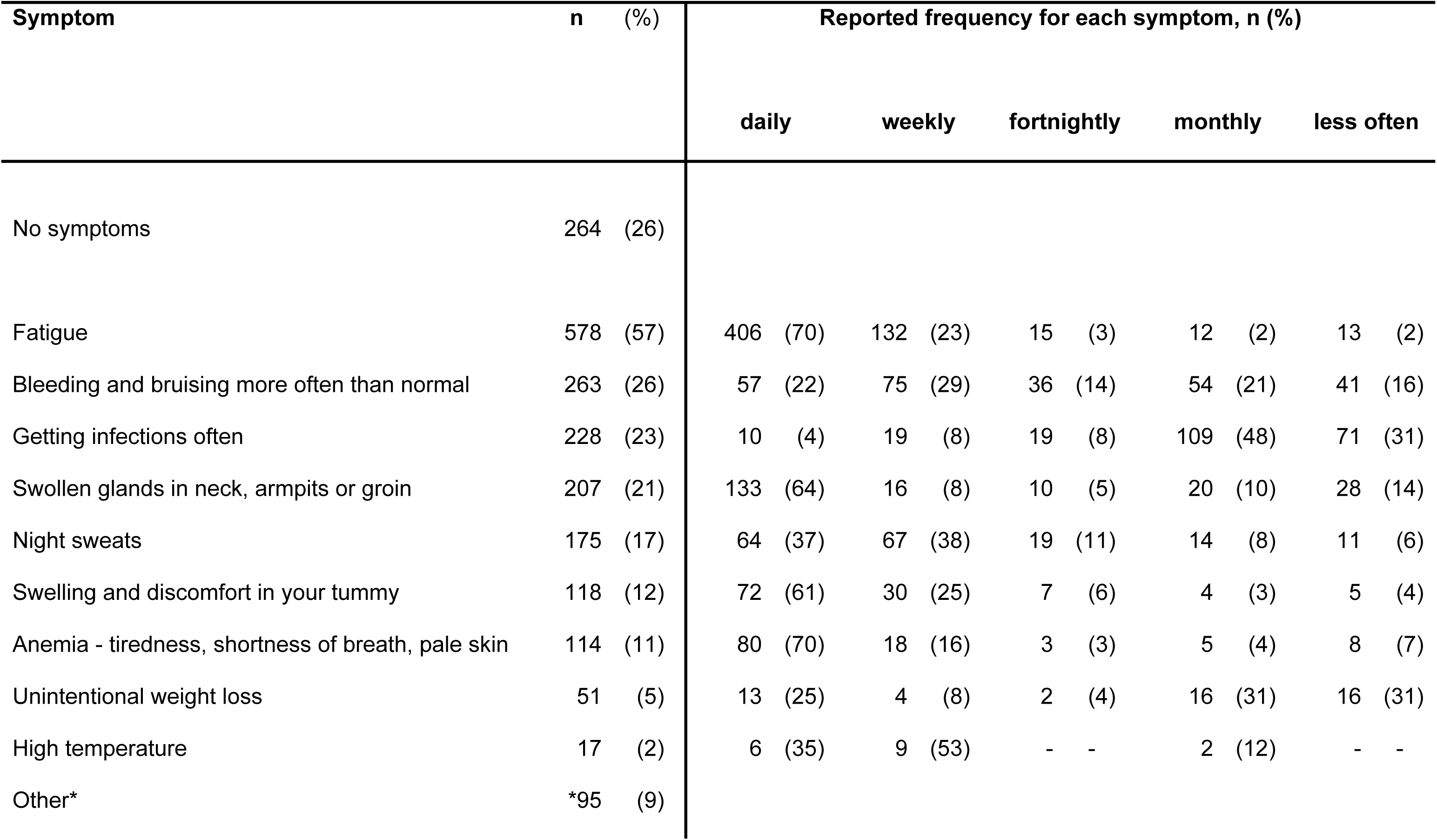
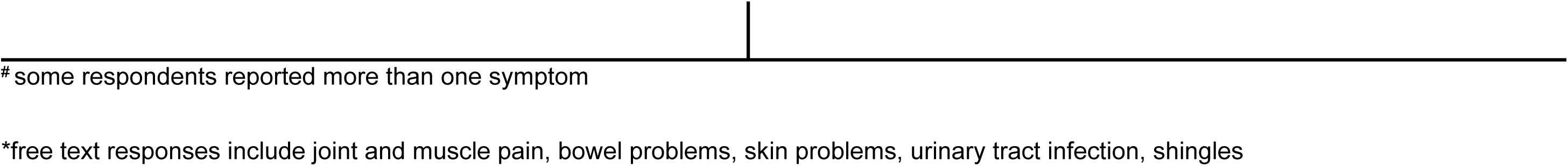
Current signs and symptoms of CLL / SLL or side effects from treatment for the 1,009 respondents^#,^ and reported frequency for each.

Respondents were asked how much their diagnosis of CLL or SLL affected their quality of life before the COVID-19 pandemic (if applicable) and ‘today’. From both an emotional and a physical perspective, the diagnosis of CLL or SLL has a higher impact now than before the pandemic (figure 3).

**Figure 3.**
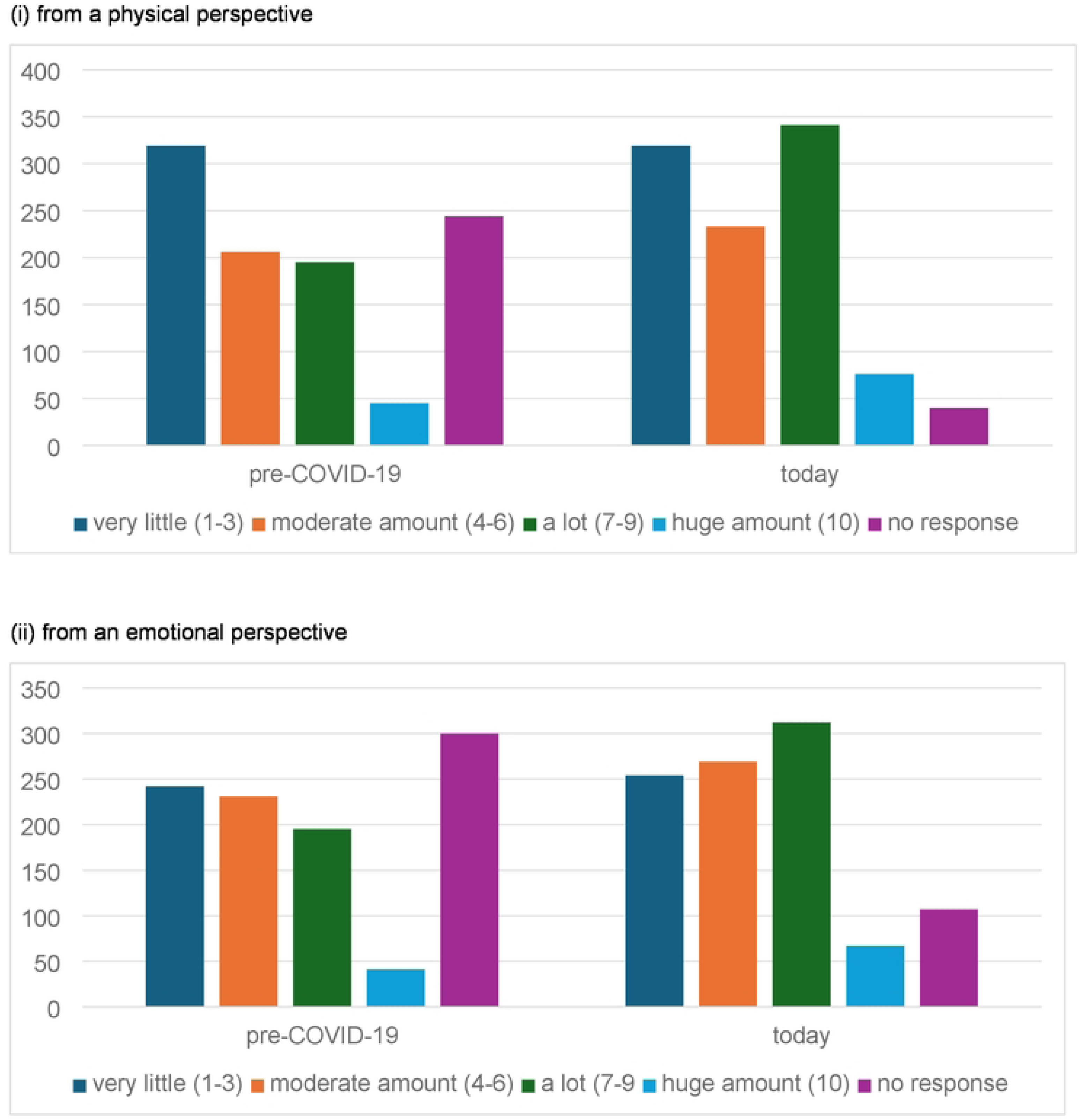
How much has your diagnosis of CLL / SLL affected your quality of life? (scale with 1= not at all, 10= huge amount)

Unsurprisingly, most respondents were currently receiving their treatment or monitoring at a hospital (829, 82%) (data not shown). Of these, almost a third said their contact was ‘mostly / all remote’ (253/829, 31%). In total, 451 (45%) of respondents were currently either on treatment or in remission. The most common treatment (at any time since diagnosis) was fludarabine, cyclophosphamide, rituximab (FCR) (table 3). This use of chemotherapy reflects that many respondents were diagnosed when these were standard therapy. Of the more modern targeted therapies, acalabrutinib, ibrutinib and venetoclax (alone or in combination with another treatment) were the most widely used. Of those currently on treatment, most (224/261, 86%) said they ‘always’ took their medication as advised, the remainder largely reported they usually took it as advised (31, 12% rated 7-9 on 10 point scale; 6 no response). Some respondents said they had accidentally missed a dose in the last four weeks (48, 18%), and a few (18, 7%) had decided to skip one or more doses (data not shown).

**Table 3.**
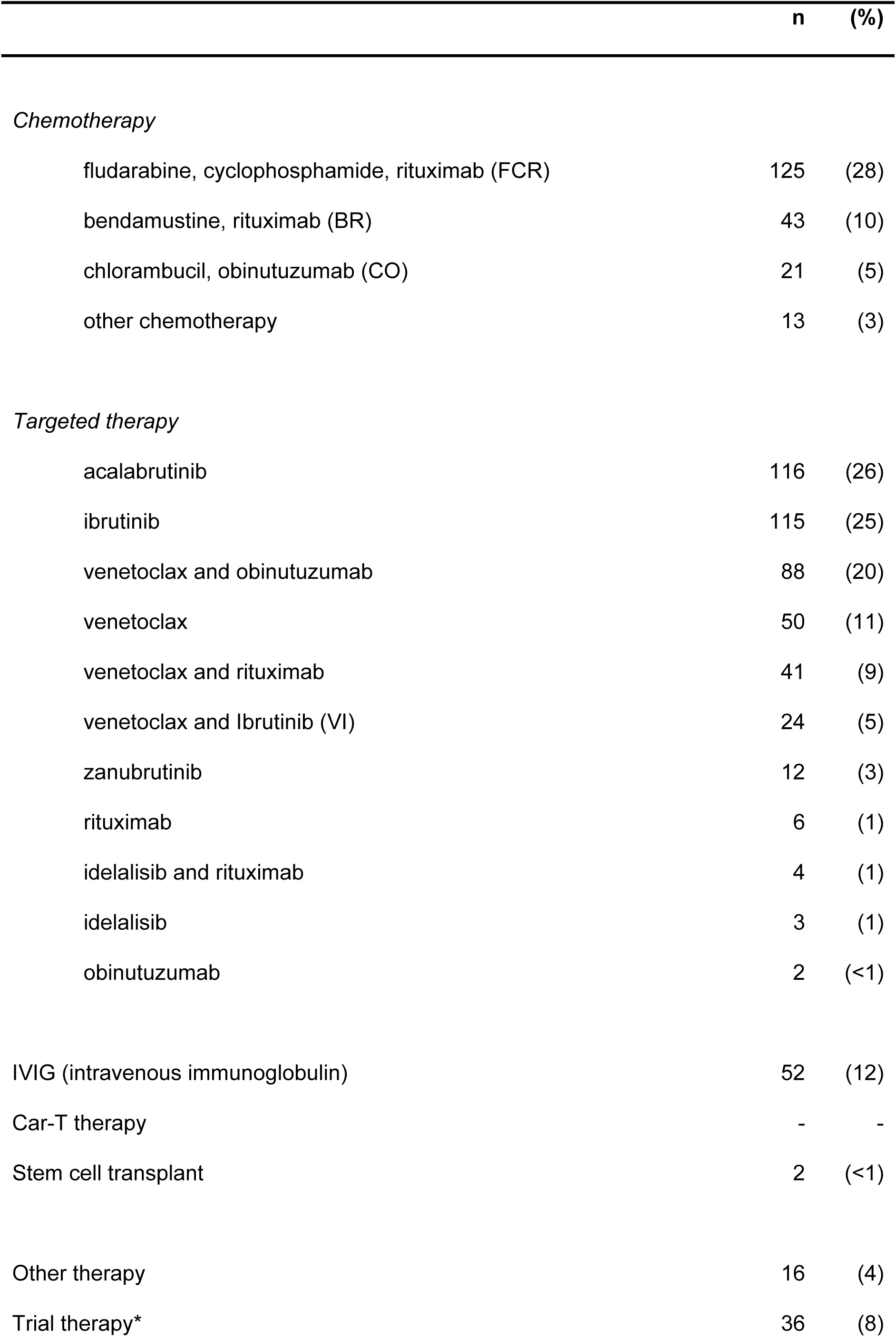
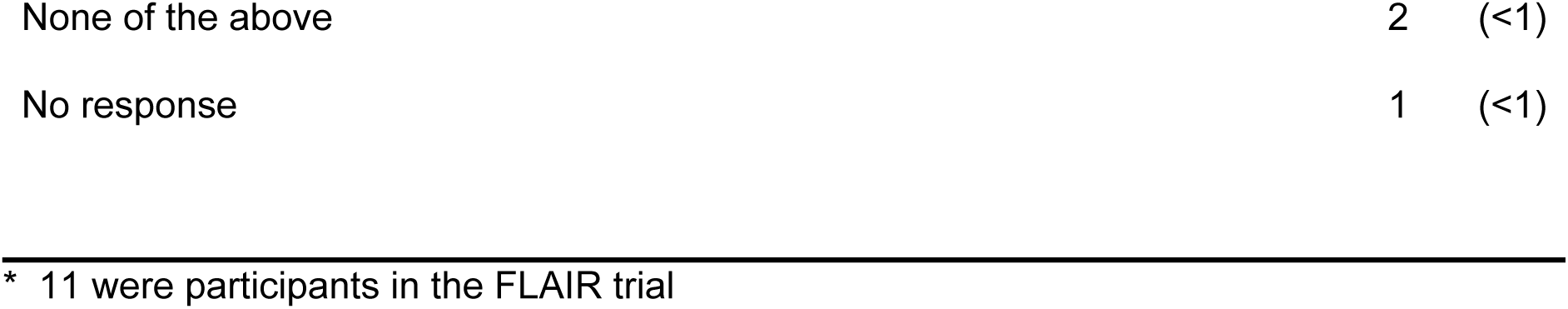
For those receiving treatment or in remission, what treatments have you had (n=451?

Respondents were asked to rate whether they felt well enough informed about treatment for CLL/SLL (on a scale of 1-10, where 1=not at all informed and 10=very well informed). Most responded they felt either well informed (7-9) (525, 52%) or very well informed (10) (205, 20%). A few felt poorly informed (1-3) (83, 8%) and almost one in five said they were moderately informed (4-6) (193, 19%) (data not shown, 3 with no response).

Almost all respondents had received a flu vaccine in the past year (918, 91%), and most had had a COVID-19 vaccination more than 9 months ago (780, 77%) and/or in the past 9 months (378, 37%). Over half the respondents had received the Shingrix vaccination (549, 54%). Just under half had received Prevenar 13 (475, 47%), and 268 (27%) had Pneumovax 2 months after Prevenar 13.

## DISCUSSION

This survey with data on more than a thousand people with CLL provides insight into the experience of living with CLL in the UK. The most striking finding is that three-quarters of those who completed our survey experience signs and symptoms of CLL or side effects from treatment for their CLL. More than half the respondents reported they experienced fatigue, and this was by far the most common problem. For those with fatigue it also had a substantial impact on their day-to-day life. For most people, their diagnosis of CLL had impacted on their quality of life from both a physical and an emotional perspective.

Almost two thirds of respondents reported their diagnosis followed a routine GP visit for some other heath condition, and was therefore likely to be unexpected. Despite this, satisfaction was high with the speed of diagnosis, explanation of their CLL diagnosis, and explanation of the next steps in their treatment.

A key strength of our survey is that it was designed by people with CLL. Hence the questions were patient focussed, addressing issues relevant to people with CLL, and were easily understandable by respondents.

As an open internet-based survey, however, our results are not necessarily representative of the UK CLL population. Those who completed our survey tended to be younger at diagnosis and were more likely to be women than the general population with CLL. Key factors contributing to these differences are that those who use the internet may not be typical of the general CLL population, and that people who volunteer to complete surveys may also not be representative. For example, both age and gender appear to influence willingness to respond to online surveys with higher response rates for younger people and for women.[11] Although our respondents almost all reported their ethnicity as white, this seems representative of the UK CLL population.[12] Nevertheless, we achieved broad geographical coverage across the UK, and across a representative range of CLL, with responses from those in active monitoring, on treatment and in remission. We also had responses from those with a recent diagnosis through to those diagnosed 30 years ago.

Other recent surveys of people with CLL have been limited by factors such as small sample size,[13] combining responses across a range of countries with different populations and healthcare systems,[13–15] and included respondents with other types of leukemia.[14,15] One survey was conducted in 12 countries, but had only 377 complete responses.[13] These respondents were predominantly from tertiary and university hospitals. In contrast to our study, demographic information was not collected so respondents could not be compared to a general population of people with CLL. Another survey was conducted across 76 countries, with 2,646 respondents of whom less than half (1,202, 45%) had CLL.[15] As in our study, fatigue was a common problem, reported by a similar proportion of respondents. It was the most frequent symptom before diagnosis (reported by 39%), and the most common side effect of treatment (reported by 44%).

There have been relatively few surveys of the experiences of people with CLL. The data presented here offer insight into the experience of living with CLL in the UK. This information will be valuable for clinicians, for health care providers and for informing future research. Future surveys should aim to have a more representative sample of the population with CLL. Strategies to facilitate this might include advertising participation through a range of blood cancer charities accessed by the CLL community, providing paper-based information about the survey and how to access it through hospital clinics, and raising awareness via as wide a range of media as possible before the survey opens. Nevertheless, our data strongly support the need for further research to help understand fatigue, its causes and effects, and how these could be mitigated. Further research would also improve our understanding of quality of life for people with CLL, what influences this, how to measure it and how could it be improved.

## Data Availability

The data are largely contained within the manuscript. As a small volunteer led charity we do not have resources to make the full data set available. If the paper is accepted, we would be happy to consider further how to make the data more avaiable

## Author contributions

The original questionnaire in 2022 was developed by Marc Auckland, then chair of the board of Trustees and the CLL Support Trustees at that time, with input from Bee Laird. The questionnaire was adapted by HL and DY for this 2024 survey. Survey data were managed by Keith Miller at Research Interactive Ltd. Data analysis was planned by DY and LD, and conducted by DY. The paper was drafted by LD with input from DY and HL. All authors agreed the final version.

## Acknowledgements

Our thanks to everyone who completed our questionnaire. Thanks also to Marc Auckland and the CLL Support Trustees for developing the 2022 questionnaire, and to Research Interactive for hosting and scripting the survey.

## Data availability statement

Data can be requested from CLL Support.

